# Resistant tuberculosis: Factors associated with unsuccessful treatment outcomes in Loreto, Peru, 2015-2023

**DOI:** 10.1101/2025.08.27.25334563

**Authors:** Evelyn Genthell Cordova-Pisco, Tery Vasquez-Hassinger, Estela Alejandra Huamán-Angeles, Karine Zevallos-Villegas, George Obregon

**Author notes:** Corresponding author: (TVH).

## Abstract

**Background:** Drug-resistant tuberculosis (DR-TB) remains a major challenge for TB control, with treatment success rates in Loreto, Peru, consistently below national averages. Nearly half of DR-TB patients in this Amazonian region experience loss to follow-up, treatment failure, or death, underscoring the need to identify region-specific factors associated with unsuccessful outcomes.

**Methods:** Retrospective cohort study including 417 MDR-TB cases registered in the national TB system in Loreto between 2015 and 2023. The unsuccessful treatment outcomes analyzed included loss to follow-up, mortality and therapeutic failure. Binary and multivariate logistic regression analyses were performed to identify risk factors associated with unsuccessful outcomes.

**Results:** Of the total number of cases, 49.5% had unsuccessful outcomes: 34.1% due to treatment dropout, 3.6% treatment failure, and 11.8% mortality. In the multivariate analysis, the factors significantly associated with unsuccessful outcomes were age between 18 and 30 years (OR: 2.8; CI: 1.1-7.1), alcoholism (OR: 3.76; CI: 1.1-12.7), drug addiction (OR: 4.48; CI: 1.2-17.2) and MDR-TB (OR: 2.2; CI: 1.2-3.8).

**Conclusions:** The identified risk factors should be the focus of priority programmatic interventions and future research aimed at optimizing regional strategies for TB control in Loreto.

## Introduction

Drug-resistant tuberculosis (DR-TB) is a major global public health problem. According to the 2024 Global Tuberculosis Report of the World Health Organization (WHO), a total of 188,666 cases of DR-TB were reported (1), representing a worrisome threat to achieving the tuberculosis control targets set for the end of this decade. The WHO End TB Strategy aims for an 80% reduction in incidence and a 90% reduction in TB mortality by 2030, compared with 2015 levels (1). However, global progress toward these goals is hindered by the growing burden of DR-TB, particularly in high-burden countries such as Peru.

At a national level, between 2015 and 2023, Peru reported 272,471 cases of TB, with a mortality rate of 5.5 per 100,000 population in 2023. Cases of DR-TB have shown an upward trend, particularly during the post-pandemic period, with treatment success rates of 78.8% for multidrug-resistant TB and rifampicin-resistant TB (RR-TB) in 2022(2). The Amazon region of Loreto represents a particular concern: treatment success rates for MDR/RR-TB were only 55.7% in 2015 and 53.8% in 2022 (2), both below the national average. This situation highlights the substantial number of patients who are not adequately diagnosed or successfully treated, thereby perpetuating TB transmission within households and communities.

Loreto accounts for 3.4% of the national MDR/RR-TB burden and is characterized by unique sociodemographic and geographic conditions: 43.5% of its population lives in extreme poverty (3), geographic accessibility is limited to air or river routes, and access to health services is deficient (4). Risk factors contributing to unsuccessful treatment outcomes in this region have been scarcely evaluated, yet they are critical to achieving cure, preventing transmission, and, most importantly, curbing the rise of resistance to anti-TB drugs.

Several studies have identified factors associated with unfavorable treatment outcomes, including age, sex, TB-HIV coinfection, TB-diabetes mellitus (DM) comorbidity, high bacillary load, and alcohol and/or drug use (5–7). These determinants reflect social and clinical conditions that continue to pose major challenges for the national health system and must be addressed with greater emphasis to improve treatment adherence and achieve higher success rates in Amazonian contexts.

To the best of our knowledge, no study has evaluated such a large cohort of DR-TB patients in the Peruvian Amazon to identify the specific factors associated with unsuccessful outcomes in Loreto, Peru, during the period 2015–2023. The findings of this study aim to contribute to the development of more effective programmatic interventions tailored to the Amazonian context.

## Methods

### Study design

This was a retrospective cohort study of patients with a confirmed diagnosis of DR-TB, using secondary data obtained from the Tuberculosis Management Information System (Spanish acronym, SIGTB) of the Regional Health Strategy for the Prevention and Control of Tuberculosis in Loreto (Spanish acronym, ESR PCTB). This system is a mandatory reporting platform covering all levels of care and for the follow-up of TB cases (8).

### Population and sample

The study population included 433 patients with DR-TB (including isoniazid-resistant TB, RR-TB, MDR-TB, and other resistance profiles) who initiated treatment and were registered in the SIGTB during the period 2015–2023 in the Loreto region. The final sample consisted of 417 patients with complete records on treatment outcomes as well as relevant sociodemographic and epidemiological variables.

### Procedure and data collection

Data were extracted from the SIGTB database, with authorization from the Regional Health Directorate of Loreto (GERESA Loreto). The database did not include personally identifiable information, thereby ensuring patient confidentiality. Data were organized into a collection form using Microsoft Excel® (v18.0), following rigorous process of cleaning, standardization, and quality control procedures. Only variables relevant to the analysis were included and coded prior to importation into statistical software for analysis. This study may be subject to selection bias due to exclusion of incomplete records, and information bias due to reliance on secondary database entries. However, standardized data collection in SIGTB likely reduced variability across centers.

### Variables

The dependent variable was treatment outcome, classified according to WHO criteria as successful (cure or treatment completed) or unsuccessful (loss to follow-up, failure, or death) (9). The independent variables included: age (categorized), sex, TB-HIV coinfection, TB-diabetes mellitus (DM) comorbidity, history of previous treatment, delay in treatment initiation, disease site (pulmonary/extrapulmonary), bacillary load, alcohol and drug use, type of drug resistance, and district of residence.

Operational definitions of the variables followed Technical Health Standard No. 104 of the Ministry of Health of Peru, consistent with WHO guidelines in effect during the study period (10).

### Statistical analysis

Statistical analyses were performed using Stata SE version 19 (StataCorp, College Station, TX, USA). Patient characteristics were summarized through descriptive analysis using descriptive statistics (frequencies). To explore the association between independent variables and unsuccessful treatment outcomes, bivariate and multivariate logistic regression models were applied to calculate crude and adjusted odds ratios (ORs) with 95% confidence intervals (95% CI). A p-value of <0.05 was considered statistically significant.

### Ethical considerations

The study protocol was approved by the Research Ethics Committee of the Regional Hospital of Loreto (No. ID-058-CIEI-2025). Authorization for access to and use of the SIGTB database was granted by the Regional Health Directorate (GERESA) of Loreto. Due to the retrospective nature of the study and the exclusive use of anonymized secondary data, informed consent from patients was not required. Confidentiality and data security were ensured throughout the entire research process.

## Results

### Characteristics of the study population

During the study period, 433 patients with a confirmed diagnosis of DR-TB from the Loreto region were registered in the SIGTB. Sixteen patients (3.7%) were excluded because their treatment outcome was not recorded, resulting in a final sample of 417 patients included in the analysis.

The mean age of the cohort was 38.1 years (IQR: 27–53 years), with the 31–45 years age group representing the largest proportion (27.1%). Male patients predominated in the sample (63.1%). Coinfection with human immunodeficiency virus (HIV) and diabetes mellitus (DM) was absent in 84.4% and 76.5% of cases, respectively. Regarding treatment history, 54.4% of patients entered the TB program as new cases, followed by 22.3% as treatment after default, 16.1% as relapses, and 7.2% as treatment failures. A total of 86.3% of patients initiated treatment within 14 days of diagnosis. Pulmonary TB was the most frequent presentation (97.1%). Negative smear microscopy at baseline was observed in 32.4% of cases. Approximately 5% of patients reported alcoholism or drug addiction. The most prevalent resistance pattern was MDR-TB (36.2%). Most patients (85.1%) came from the district of Maynas city (Table 1).

**Table 1.** Clinical and sociodemographic characteristics of patients with drug-resistant TB in Loreto, 2015–2023.

| Variables |  | n | % |
| --- | --- | --- | --- |
| Age | < 18 years | 34 | 8.2 |
|  | 18 - 30 years | 93 | 22.3 |
|  | 31 - 45 years | 113 | 27.1 |
|  | 46 - 60 years | 109 | 26.1 |
|  | > 60 years | 68 | 16.3 |
| Sex | Female | 154 | 36.9 |
|  | Male | 263 | 63.1 |
| TB-HIV coinfection | No | 352 | 84.4 |
|  | Yes | 43 | 10.3 |
|  | Not evaluated | 22 | 5.3 |
| TB-DM | No | 319 | 76.5 |
|  | Yes | 76 | 18.2 |
|  | Not evaluated | 22 | 5.3 |
| Previous treatment history | New case | 227 | 54.4 |
|  | Relapse | 67 | 16.1 |
|  | Treatment after loss to follow-up | 93 | 22.3 |
|  | Failure | 30 | 7.2 |
| Delay in treatment initiation | < 14 days | 360 | 86.3 |
|  | > 14 days | 50 | 12.0 |
|  | Missing data | 7 | 1.7 |
| TB location | Pulmonary | 405 | 97.1 |
|  | Extrapulmonary | 12 | 2.9 |
| Bacillary load | Negative | 135 | 32.4 |
|  | Paucibacilar | 2 | 0.5 |
|  | + | 121 | 29.0 |
|  | ++ | 75 | 18.0 |
|  | +++ | 60 | 14.4 |
|  | Not performed | 24 | 5.8 |
| Alcoholism | No | 396 | 95.0 |
|  | Yes | 21 | 5.0 |
| Drug addiction | No | 395 | 94.7 |
|  | Yes | 22 | 5.3 |
| Resistance | Other | 119 | 28.5 |
|  | TB-H | 67 | 16.1 |
|  | TB-R | 75 | 18.0 |
|  | TB-Z | 1 | 0.2 |
|  | TB-Fluoroquinolones | 2 | 0.5 |
|  | MDR-TB | 151 | 36.2 |
|  | Pre-XDR TB | 2 | 0.5 |
| District | Periphery | 62 | 14.9 |
|  | Maynas city | 355 | 85.1 |
|  | **Total** | **417** |  |

### Treatment Outcomes

Of the 417 patients analyzed, 50.6% (211) achieved treatment success, while 49.4% (206) experienced unsuccessful outcomes. Within the latter group, loss to follow-up (34%) was the most frequent cause, followed by mortality (11.8%) and therapeutic failure (3.6%) (Figure I).

**Figure 1.**
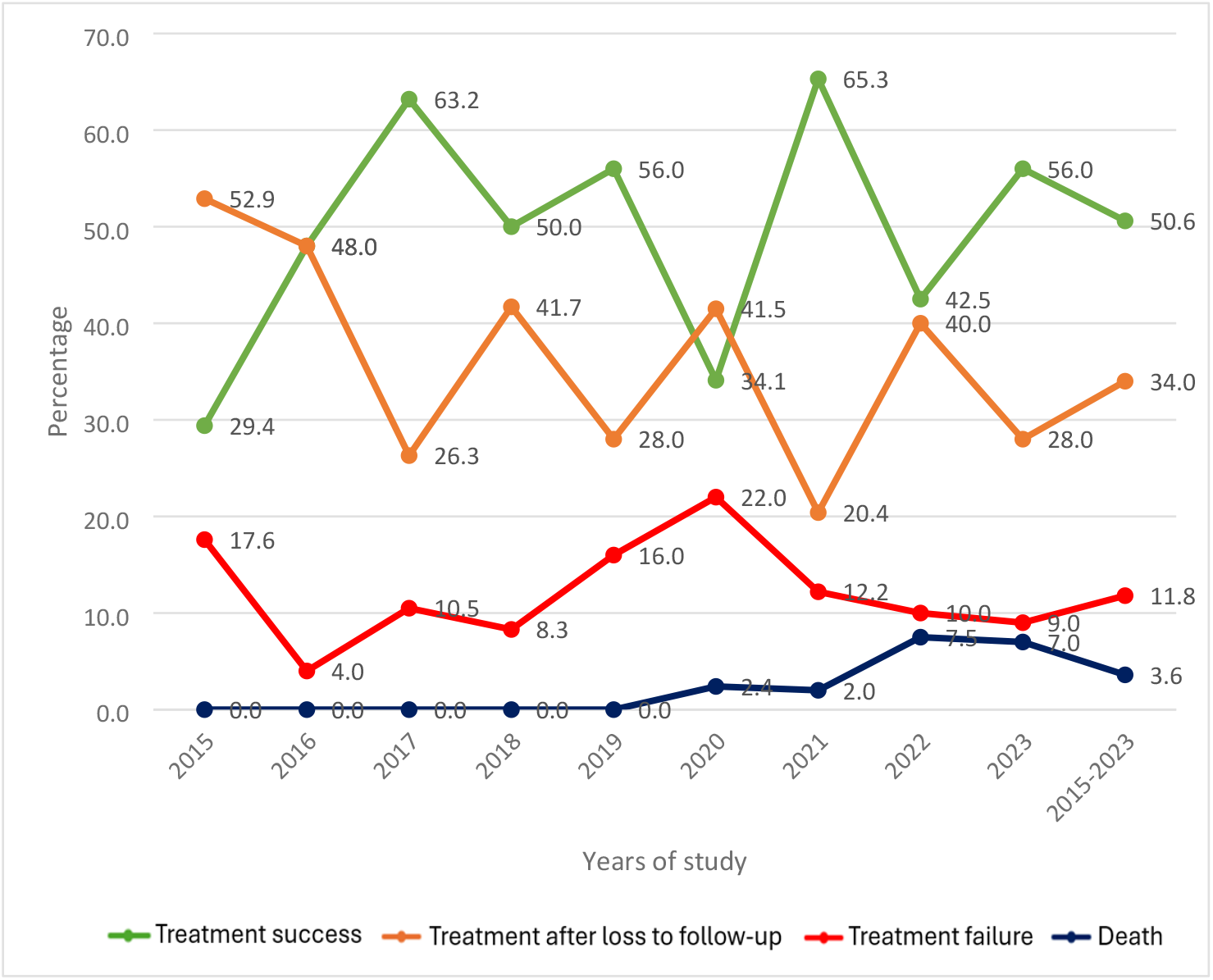
Proportion of treatment outcomes in patients with DR-TB in Loreto, 2015–2023.

Bivariate logistic regression showed significant associations between unsuccessful treatment outcomes and the following variables: age 18–30 years (OR: 3.7; CI: 1.6–8.5; p=0.002); TB-HIV coinfection (OR: 2.4; CI: 1.2–4.7; p=0.012); history of treatment after default (“treatment after loss to follow-up”) (OR: 2.9; CI: 1.6–5.2; p<0.001); high bacillary load (+++) (OR: 1.9; CI: 1.0–3.5; p=0.041); alcoholism (OR: 4.6; CI: 1.5–14.1; p=0.006); drug addiction (OR: 7.0; CI: 2.0–24.2; p=0.002); and MDR-TB resistance (OR: 1.9; CI: 1.1–3.1; p=0.011).

According to the multivariate analysis, treatment outcomes were significantly affected among patients aged 18–30 years, who were 3.1 times more likely to experience an unsuccessful outcome (CI: 1.1–8.1; p=0.023). Significant associations were also found with a history of alcoholism (OR: 3.6; CI: 1.1–12.2; p=0.039), drug addiction (OR: 4.3; CI: 1.1–16.6; p=0.032), and MDR-TB (OR: 2.2; CI: 1.2–3.8; p=0.006), all of which were independently associated with unsuccessful treatment outcomes (Table 2).

**Table 2.** Factors Associated with Unsuccessful Outcomes in Patients with Drug-Resistant TB in Loreto, 2015–2023.

| Predictors |  |  | Bivariate analysis |  | Multivariate analysis |  |
| --- | --- | --- | --- | --- | --- | --- |
|  | Success<br>n (%) | Unsuccessful<br>n (%) | OR (95% CI) | p-value | aOR (95% CI) | p-value |
| **Age** |  |  |  |  |  |  |
| < 18 years | 21 (5.0) | 13 (3.1) | 1 |  | 1 |  |
| 18 - 30 years | 28 (6.7) | 65 (15.6) | **3.7 (1.6-8.5)** | **0.002** | **3.1 (1.1-8.1)** | **0.023** |
| 31 - 45 years | 58 (13.9) | 55 (13.2) | 1.5 (0.7-3.3) | 0.286 | 1.2 (0.4-3.0) | 0.706 |
| 46 - 60 years | 59 (14.1) | 50 (12.0) | 1.4 (0.6-3.0) | 0.435 | 1.1 (0.4-2.7) | 0.918 |
| > 60 years | 45 (10.8) | 23 (5.5) | 0.8 (0.3-1.9) | 0.660 | 0.8 (0.3-2.2) | 0.684 |
| **Sex** |  |  |  |  |  |  |
| Female | 86 (20.6) | 68 (16.3) | 1 |  | 1 |  |
| Male | 125 (30.0) | 138 (33.1) | 1.4 (0.9-2.1) | 0.102 | 1.2 (0.7-1.9) | 0.526 |
| **TB-HIV coinfection** |  |  |  |  |  |  |
| No | 188 (45.1) | 164 (39.3) | 1 |  | 1 |  |
| Yes | 14 (3.4) | 29 (7.0) | **2.4 (1.2-4.7)** | **0.012** | 1.23 (0.5-2.8) | 0.609 |
| Not evaluated | 9 (2.2) | 13 (3.1) | - |  | - |  |
| **TB-DM comorbidity** |  |  |  |  |  |  |
| No | 159 (38.1) | 160 (38.4) | 1 |  | 1 |  |
| Si | 44 (10.6) | 32 (7.7) | 0.7 (0.4-1.2) | 0.208 | 0.9 (0.5-1.6) | 0.617 |
| Not evaluated | 8 (1.9) | 14 (3.4) | - |  | - |  |
| **Previous treatment history** |  |  |  |  |  |  |
| New case | 130 (31.2) | 97 (23.3) | 1 |  | 1 |  |
| Relapse | 14 (3.4) | 16 (3.8) | 1.5 (0.7-3.3) | 0.274 | 1.2 (0.5-2.9) | 0.697 |
| Treatment after loss to follow-up | 21 (5.0) | 46 (11.0) | **2.9 (1.6-5.2)** | **0.000** | 1.5 (0.7-3.1) | 0.181 |
| Failure | 46 (11.0) | 47 (11.3) | 1.4 (0.8-2.2) | 0.203 | 1.2 (0.7-2.1) | 0.599 |
| **Delay in treatment initiation** |  |  |  |  |  |  |
| < 14 days | 182 (43.6) | 178 (42.7) | 1 |  | 1 |  |
| > 14 days | 29 (7.0) | 21 (5.0) | 0.7 (0.4-1.3) | 0.325 | 0.7 (0.3-1.5) | 0.363 |
| Missing data | 0 | 7 (1.7) | - |  | - |  |
| **TB location** |  |  |  |  |  |  |
| Pulmonary | 208 (49.9) | 197 (47.2) | 1 |  | 1 |  |
| Extrapulmonary | 3 (0.7) | 9 (2.2) | 3.2 (0.8-11.9) | 0.087 | 3.7 (0.8-16.1) | 0.085 |
| **Bacillary load** |  |  |  |  |  |  |
| Negative | 80 (19.2) | 55 (13.2) | 1 |  | 1 |  |
| Paucibacilar | 1 (0.2) | 1 (0.2) | 1.4 (0.1-23.8) | 0.793 | 2.0 (0.0-127.8) | 0.751 |
| + | 61 (14.6) | 60 (14.4) | 1.4 (0.9-2.3) | 0.156 | 1.3 (0.7-2.4) | 0.333 |
| ++ | 34 (8.2) | 41 (9.8) | 1.7 (0.9-3.1) | 0.053 | 1.6 (0.8-3.2) | 0.163 |
| +++ | 26 (6.2) | 34 (8.2) | **1.9 (1.0-3.5)** | **0.041** | 1.6 (0.8-3.3) | 0.203 |
| Not performed | 9 (2.2) | 15 (3.6) | - |  | - |  |
| **Alcoholism** |  |  |  |  |  |  |
| No | 207 (49.6) | 189 (45.3) | 1 |  | 1 |  |
| Yes | 4 (1.0) | 17 (4.1) | **4.6 (1.5-14.1)** | **0.006** | **3.6 (1.1-12.2)** | **0.039** |
| **Drug addiction** |  |  |  |  |  |  |
| No | 208 (49.9) | 187 (44.8) | 1 |  | 1 |  |
| Yes | 3 (0.7) | 19 (4.6) | **7.0 (2.0-24.2)** | **0.002** | **4.3 (1.1-16.6)** | **0.032** |
| **Resistance** |  |  |  |  |  |  |
| Other | 61 (14.6) | 58 (13.9) | 1 |  | 1 |  |
| TB-H | 51 (12.2) | 16 (3.8) | 0.3 (0.2-0.6) | 0.001 | 0.4 (0.2-0.9) | 0.023 |
| TB-R | 44 (10.6) | 31 (7.4) | 0.7 (0.4-1.3) | 0.314 | 0.8 (0.4-1.6) | 0.530 |
| TB-Z | 0 | 1 (0.2) | - |  | - |  |
| TB-Fluoroquinolones | 1 (0.2) | 1 (0.2) | 1.0 (0.1-17.2) | 0.972 | 1.6 (0.1-29.2) | 0.730 |
| MDR-TB | 54 (12.9) | 97 (23.3) | **1.9 (1.1-3.1)** | **0.011** | **2.2 (1.2-3.8)** | **0.006** |
| Pre-XDR TB | 0 | 2 (0.5) | - |  | - |  |
| **District** |  |  |  |  |  |  |
| Periphery | 31 (7.4) | 30 (7.2) | 1 |  | 1 |  |
| Maynas city | 180 (43.2) | 176 (42.2) | 1.3 (0.7-2.3) | 0.319 | 1.7 (0.8-3.2) | 0.120 |

## Discussion

This study reveals that approximately half (49.4%) of patients with DR-TB treated in the Loreto region between 2015 and 2023 experienced unsuccessful treatment outcomes (treatment loss to follow-up, failure, or mortality), a figure far below the target set at the national level (11). Among the main factors associated with unsuccessful outcomes identified in our multivariate analysis were age range 18–30 years, alcoholism, drug addiction, and MDR-TB. These findings highlight the urgent need to strengthen TB control strategies in Amazonian settings characterized by high social and geographic vulnerability.

Young adults (18–30 years) constitute a particularly vulnerable population to unfavorable therapeutic outcomes in TB management (12). A possible explanation lies in the demands associated with active working life, as they often engage in informal or unstable employment, which limits their availability to consistently attend health services. This situation may be further aggravated by the need to prioritize economic subsistence over treatment continuity, especially in contexts of poverty or limited institutional support. In addition, behavioral and social factors may play a role, as this group tends to have greater mobility and engage in social practices that may involve psychoactive substance use and increased community exposure—factors that not only heighten the risk of acquiring the disease but also of failing to adhere adequately to treatment once diagnosed (13–18).

Alcohol and drug use emerged as key determinants associated with unsuccessful therapeutic outcomes, with up to a fourfold higher risk compared to patients without these behaviors. These findings are consistent with previous studies, which reported that illicit drug use is associated with a twofold increase in the risk of unfavorable outcomes, such as loss to follow-up, among patients with DR-TB (19)(20). Alcohol consumption exerts a multifactorial negative impact, being linked to a higher probability of loss to follow-up, more contagious clinical forms such as cavitary tuberculosis, and impaired immune response against *Mycobacterium tuberculosis*, all of which compromise the effectiveness of the pharmacological regimen (21). In addition, these behaviors impair cognitive function, making it more difficult to sustain adherence to the prescribed pharmacological regimen (20,22–24). At the national level, current regulations stipulate that patients who screen positive for alcohol or illicit substance dependence should be referred to the mental health unit of their corresponding health facility, and if specialized care is required, to a higher-level facility for management. However, despite the implementation of this management protocol since 2013, the rates of unsuccessful therapeutic outcomes in this group remain high. This underscores the need for a multidisciplinary approach that comprehensively addresses factors such as precarious living conditions, immune system compromise, and poor treatment adherence, in order to improve outcomes in this vulnerable population.

The presence of MDR-TB was associated with a higher risk of unfavorable outcomes, with patients being twice as likely to experience unsuccessful results. This evidence is consistent with findings from a study conducted in Lima, the Peruvian region with the highest prevalence of DR-TB cases, where patients treated for MDR-TB had a threefold higher risk of loss to follow-up compared with those with drug-susceptible TB (7). However, unlike the predominantly urban context of Lima, the Loreto region faces greater challenges due to its vast geography and limited access to health services for drug susceptibility testing, which further increases the risk of delayed DR-TB diagnosis, loss to follow-up, and, consequently, unfavorable clinical outcomes (25). These circumstances reinforce the need to adapt TB control strategies to the Amazonian context.

On the other hand, no association was found between previous treatment history and unsuccessful outcomes, possibly because more than 50% of the cases in this study were new. Nevertheless, regardless of whether the patient had a prior history of TB treatment or was a new case (23), this phenomenon reflects the circulation of resistant strains within the community, underscoring the urgent need to decentralize and strengthen drug susceptibility testing in Loreto (18).

Similarly, other studies have identified significant associations with additional factors such as TB-DM comorbidity and TB-HIV coinfection (19,20,22,26–28). These conditions are mainly linked to immune system complications and the development of other opportunistic diseases, thereby increasing the clinical burden and treatment complexity, which in turn favors TB progression and unsuccessful outcomes (29,30). The lack of association in our study may be related to the limited availability of HIV and DM screening for all patients, due to logistical issues and shortages (31), resulting in underreporting of these data in the system.

The strengths of this study include the cohort size and the use of population-based data from a region with limited geographic access to drug susceptibility testing, which provides representative evidence for dispersed and hard-to-reach populations. Nevertheless, the study is subject to limitations inherent to the use of secondary data, such as incomplete records (less than 10%), underreporting of DR-TB cases, and missing information for certain clinical and social variables of interest.

## Conclusions

The findings of this study highlight the need to design and implement comprehensive and targeted interventions that effectively address the identified risk factors, with priority given to drug susceptibility testing in the region and the evaluation of regional mental health approaches for DR-TB cases.

## Data Availability

All data underlying the findings of this study are fully available without restriction and are provided within the manuscript and its Supporting Information files

## Acknowledgments

We gratefully acknowledge the Regional Health Strategy for the Prevention and Control of Tuberculosis of the Regional Health Directorate of Loreto, and to the Research Committee of the Regional Hospital of Loreto.

